# Intracranial Atherosclerotic Disease in Acute Ischemic Stroke: Clinical Predictors, Imaging Profiles, and Treatment Outcomes: An Analysis from RITE Registry

**DOI:** 10.1101/2024.11.24.24317863

**Authors:** Fawaz F. Alotaibi, Michael Mlynash, M. Aldawsari Muteb, Nowar Z. AlAbidien, Mohammed Alqahtani, Hussain BinAmir, Ammar AlKawi, Abdulrahman A. Alreshaid, Mohamed AlZawahmah, Adel Alhazzani, Hani Alabdaly, Abdulmajeed Alanazi, Nouf Almansour, Gregory W. Albers, Ashfaq Shuaib, Fahad S. Al-Ajlan

**Author notes:** **Corresponding author:** Fahad S Al-Ajlan, MD, Neuroscience Center, King Faisal Specialist Hospital, Altakhasosy Street, P.O. Box-3354 (MBC - 76), Riyadh, Riyadh 11211, Saudi Arabia.

## Abstract

**Background:** Intracranial atherosclerotic disease (ICAD) is a leading cause of ischemic stroke worldwide, with a significant prevalence in specific populations. This study aims to compare the clinical characteristics, imaging profiles, and outcomes of stroke patients with large vessel occlusion (LVO) caused by ICAD versus non-ICAD etiologies, particularly in those transferred for endovascular treatment (EVT).

**Methods:** A retrospective analysis was conducted using data from the Riyadh Thrombectomy Registry, including 219 patients with acute ischemic stroke who underwent EVT between January 2019 and March 2024. Clinical, demographic, and imaging data were collected and compared between ICAD and non-ICAD groups. Imaging modalities included computed tomography perfusion (CTP) and angiography, with key metrics such as the hypoperfusion intensity ratio (HIR) and cerebral blood volume (CBV) index evaluated for prognostic utility.

**Results:** Patients with ICAD were older and demonstrated a higher prevalence of vascular risk factors. ICAD patients presented with better ASPECTS scores, smaller ischemic core volumes, and higher mismatch ratios, reflecting better collateral circulation. However, despite favorable imaging profiles, ICAD patients exhibited higher mortality rates, particularly in cases involving basilar artery occlusion. HIR and CBV indices demonstrated modest diagnostic performance in distinguishing ICAD from non-ICAD, with an area under the curve (AUC) of 0.62 for HIR and 0.57 for CBV.

**Conclusions:** ICAD patients undergoing EVT showed distinct clinical and imaging characteristics compared to non-ICAD patients. Despite better collateral circulation, ICAD is associated with higher mortality, especially in posterior circulation strokes. Further research is required to optimize EVT strategies and explore adjunctive therapies tailored to this challenging patient population.

## Introduction

Intracranial atherosclerotic disease (ICAD) is a major cause of ischemic stroke worldwide. ^(1)^ The prevalence of ICAD is estimated to be 30–50% of acute ischemic strokes (AIS) in Asian populations, 15–29% in Black populations, and 5–10% in White populations. ^(2)^ A single-center, hospital-based retrospective study in Saudi Arabia enrolling patients with ICAD-related AIS between 2017 and 2020 found that 26% of 512 AIS cases were attributable to ICAD. ^(3)^ Similarly, another study from the southern region of Saudi Arabia, which included 201 stroke patients, reported that 45.77% had intracranial stenosis. Significant risk factors identified included hypertension, ischemic heart disease, obesity, and smoking, with obesity showing the strongest association. Among those treated, 72% experienced no stroke recurrence during follow-up. ^(4)^ The incidence of AIS caused by ICAD is projected to increase with population growth in regions where ICAD is prevalent. ^(5)^

Perfusion imaging using artificial intelligence (AI) software has emerged as a key tool for the preoperative evaluation of mechanical thrombectomy (MT). This approach automatically processes images derived from computed tomography perfusion (CTP) systems, enabling the identification and quantification of the infarct core and ischemic penumbra. ^(6)^ Parameters of perfusion imaging, such as the hypoperfusion intensity ratio (HIR), are valuable in assessing collateral flow in patients with anterior large vessel occlusion (LVO). Both the HIR and cerebral blood volume (CBV) index are associated with underlying ICAD and may serve as predictors of ICAD before endovascular treatment (EVT). ^(7)^

Significant advancements have been made in the treatment of acute strokes secondary to LVO. EVT is highly effective when performed within 24 hours of symptom onset in appropriately selected patients. ^(8)^ However, because EVT facilities are not widely available, many patients require inter-hospital transfer for treatment, which increases the risk of complications. Factors associated with better outcomes include shorter ’in-out’ times at the primary center, smaller ischemic core volume, better pial collateral circulation, and lower HIR on perfusion imaging. ^(9)^ Patients with ICAD are particularly sensitive to changes in blood pressure and intracranial perfusion pressure. Recent studies have shown that ICAD patients often present with smaller ischemic core volumes and reduced hypoperfusion intensity, as indicated by a lower HIR and smaller tissue volumes with prolonged Tmax >6 seconds. ^(10)^

The primary aim of this study is to retrospectively evaluate differences in clinical characteristics, imaging features, and outcomes of stroke patients with large vessel occlusions caused by intracranial atherosclerosis compared to those without intracranial atherosclerosis. The focus is specifically on patients who were transferred to the comprehensive stroke center at King Faisal Specialist Hospital and Research Center in Riyadh.

## Methods

This study was approved by the institutional review board of King Faisal Specialist Hospital and Research Center in Riyadh, Saudi Arabia (RAC #2241329).

### Study Participants

We retrospectively analyzed 219 patients from the Riyadh Thrombectomy Registry (RITE). The inclusion criteria consisted of adults aged 18 years and older who were diagnosed with acute ischemic stroke with evidence of large or medium vessel occlusion and underwent mechanical thrombectomy, with or without an underlying etiology of ICAD, between January 2019 and March 2024.

Collected clinical data included demographic variables (age and gender) and risk factors, such as a history of hypertension, diabetes mellitus, dyslipidemia, coronary artery disease, atrial fibrillation, smoking, and use of anticoagulants. Laboratory data included hemoglobin levels, platelet counts, international normalized ratio (INR), glycosylated hemoglobin (HbA1c), and creatinine levels.

### Imaging Data

All stroke patients underwent a comprehensive CT scan, which included a non-contrast CT, CT angiography of the head and neck, and CT perfusion, using a 256-slice multi-detector CT scanner (Brilliance iCT). Initially, a non-contrast CT scan of the head was performed to rule out intracranial bleeding, followed by a CT angiogram of the head and neck, and subsequent CT perfusion imaging. Whole-brain helical NCCT (120 kVp, 100–350 auto-mAs) was performed with a 5-mm section thickness. CT perfusion parameters were acquired in a periodic spiral pattern.

A high-pressure syringe was used to inject 70–90 mL of the contrast agent iopamidol at a flow rate ranging from 4.0 to 6.0 mL/s. Subsequently, the tube was flushed with 30 mL of physiological saline, and the scan commenced after a 5-second delay. The imaging spanned from the foramen magnum to the level above the lateral ventricle, utilizing an 80-mm collimation, tube voltage of 80 kV, and tube current of 100 mA. The perfusion maps and their associated parameters were automatically generated using RAPID software (iSchemaView, Menlo Park, CA; version 5.0.4).

The ischemic core was defined as a tissue volume with cerebral blood flow of <30% on CTP imaging. Hypoperfusion was defined as a volume of tissue with a Tmax >6 seconds on CTP. The mismatch ratio was calculated by dividing the ischemic core volume by the lesion volume with a Tmax >6 seconds. The mismatch volume was calculated by subtracting the ischemic core volume from the lesion volume with a Tmax >6 seconds.

HIR (hypoperfusion intensity ratio) was defined as the ratio of the volume of the “Tmax >10 seconds” lesion to the volume of the “Tmax >6 seconds” lesion. The CBV index was calculated as the ratio of the mean CBV within the “Tmax >6 seconds” lesion in the ipsilateral hemisphere to the mean CBV of the unaffected brain area.

### Statistical Analysis

Categorical variables were compared between the ICAD and non-ICAD groups using the χ² test, while continuous and ordinal variables were analyzed using the Mann-Whitney U test. Differences in medians, along with their 95% confidence intervals, were estimated using the Hodges-Lehmann method.

Receiver operating characteristic (ROC) curve analysis was performed to differentiate between groups based on continuous variables. Optimal cut-off values were determined using Youden’s J-statistic.

Binary logistic regression analysis was conducted to assess the association of ICAD with outcomes, adjusting for imbalances between ICAD and non-ICAD groups that were significant at *p*<0.1. A backward elimination procedure was applied to retain only variables with *p*<0.1 in the final model after adjustments.

All statistical tests were considered significant at a threshold of α=0.05. Analyses were performed using SAS version 9.4 and IBM SPSS version 29.

## Results

Baseline characteristics for the two groups are presented in (Table 1). Patients with ICAD were older and had a higher prevalence of prior stroke, hypertension, diabetes mellitus, and smoking. However, they were less likely to have atrial fibrillation or be on anticoagulation therapy. There was a non-significant trend toward a lower prevalence of coronary artery disease (CAD) and lower HbA1c levels in ICAD patients (Supplementary 1).

**Table 1:** Baseline characteristics.

|  | ICAD | Non-ICAD | p-value | Difference<br>between medians<br>(95% CI) |
| --- | --- | --- | --- | --- |
|  | N=107 | N=112 |  |  |
| Age, years | 61 (54-72) | 58 (45-69) | **0.024** | 5 (1-9) |
| Sex, male | 70 (65) | 66 (59) | 0.333 |  |
| Hx CAD | 31 (29) | 45 (41) | 0.073 |  |
| Hx of Stroke | 31 (29) | 18 (16) | **0.022** |  |
| Hx DLP | 27 (26) | 25 (22) | 0.585 |  |
| Hx Afib | 5 (5) | 17 (15) | **0.010** |  |
| Hx<br>Anticoagulation | 4 (4) | 23 (21) | **<0.001** |  |
| Hx HTN | 71 (66) | 55 (49) | **0.010** |  |
| Hx DM | 64 (60) | 39 (35) | **<0.001** |  |
| Hx Smoking | 59 (55) | 36 (32) | **<0.001** |  |

The occlusion locations in intracranial arteries differed significantly between the two groups. Pairwise comparisons showed that ICAD patients had a significantly higher prevalence of basilar artery occlusions (*p*<0.001; Table 2a). ICAD patients also received more intravenous tPA prior to transfer to the tertiary center for EVT.

**Table 2a:** Clinical and Imaging Characteristics.

|  | ICAD | Non-ICAD | p-value |
| --- | --- | --- | --- |
|  | N=107 | N=112 |  |
| Multiple vessels | 56 (52) | N/A |  |
| Culprit vessel | 47 (44) | N/A |  |
| IV tPA | 32 (30) | 42 (38) | 0.235 |
| General Anesthesia | 47 (44) | 43 (38) | 0.406 |
| Occlusion location |  |  | 0.009 |
| ICA | 14 (13) | 14 (13) |  |
| M1 | 55 (51) | 63 (56) |  |
| M2-M3 | 19 (18) | 29 (26) |  |
| Tandem | 2 (2) | 2 (2) |  |
| Basilar | 12 (11) | 0 (0) |  |
| Other | 5 (5) | 4 (4) |  |

ICAD patients demonstrated better ASPECTS, smaller core lesion volumes, higher mismatch ratios, and better HIR. The size of the Tmax >10s lesions in perfusion imaging was nominally smaller in the ICAD group (indicating less severe hypoperfusion), and ICAD patients showed a slightly better CBV index (Table 2b).

**Table 2b:**
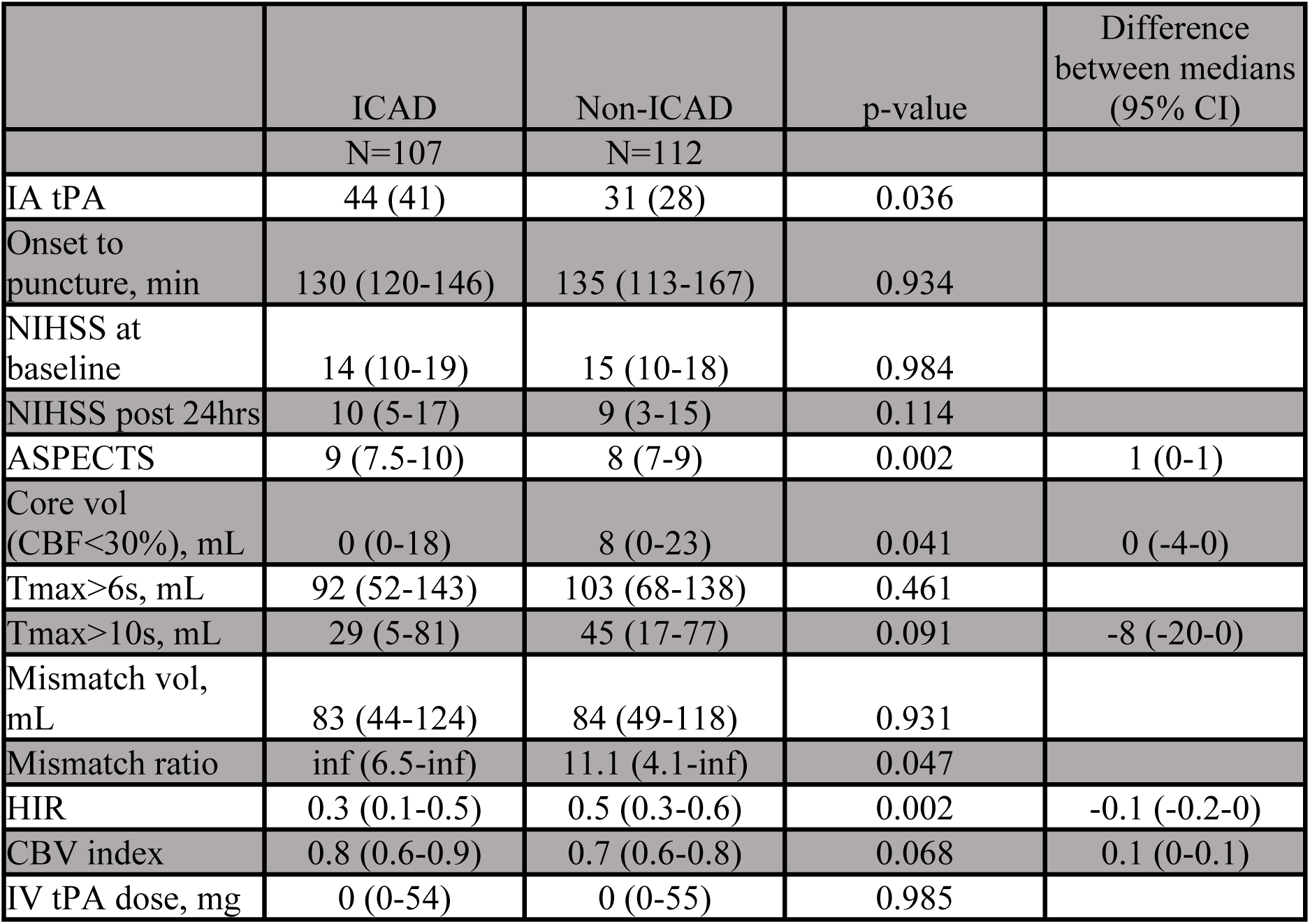
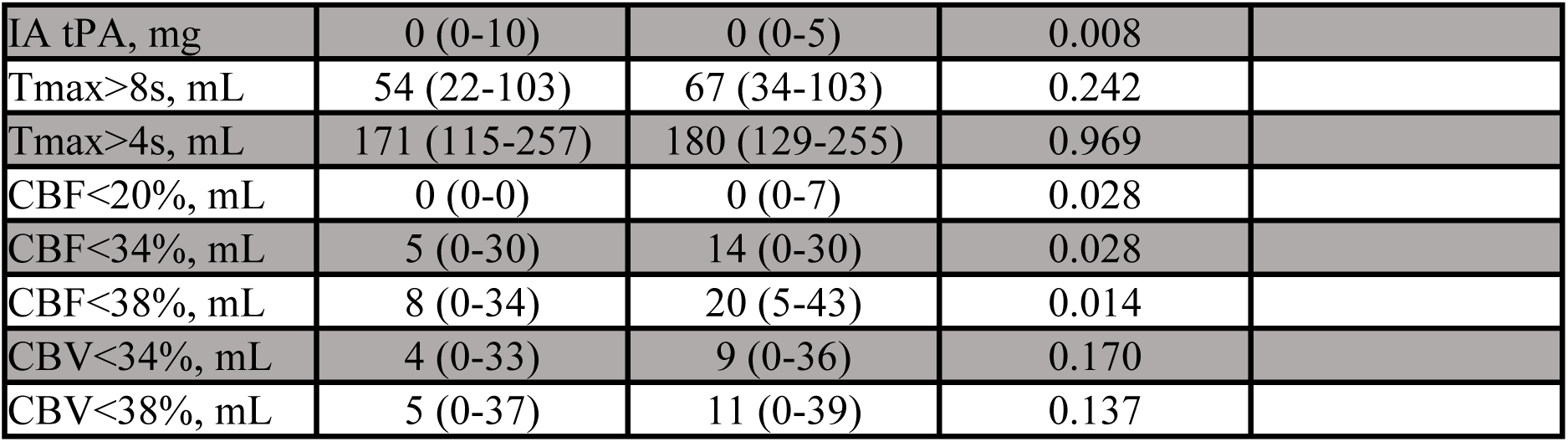
(Clinical and Imaging criteria)

|  | ICAD | Non-ICAD | p-value | Difference between medians (95% CI) |
| --- | --- | --- | --- | --- |
|  | N=107 | N=112 |  |  |
| IA tPA | 44 (41) | 31 (28) | 0.036 |  |
| Onset to puncture, min | 130 (120-146) | 135 (113-167) | 0.934 |  |
| NIHSS at baseline | 14 (10-19) | 15 (10-18) | 0.984 |  |
| NIHSS post 24hrs | 10 (5-17) | 9 (3-15) | 0.114 |  |
| ASPECTS | 9 (7.5-10) | 8 (7-9) | 0.002 | 1 (0-1) |
| Core vol (CBF<30%), mL | 0 (0-18) | 8 (0-23) | 0.041 | 0 (-4-0) |
| Tmax>6s, mL | 92 (52-143) | 103 (68-138) | 0.461 |  |
| Tmax>10s, mL | 29 (5-81) | 45 (17-77) | 0.091 | -8 (-20-0) |
| Mismatch vol, mL | 83 (44-124) | 84 (49-118) | 0.931 |  |
| Mismatch ratio | inf (6.5-inf) | 11.1 (4.1-inf) | 0.047 |  |
| HIR | 0.3 (0.1-0.5) | 0.5 (0.3-0.6) | 0.002 | -0.1 (-0.2-0) |
| CBV index | 0.8 (0.6-0.9) | 0.7 (0.6-0.8) | 0.068 | 0.1 (0-0.1) |
| IV tPA dose, mg | 0 (0-54) | 0 (0-55) | 0.985 |  |
| IA tPA, mg | 0 (0-10) | 0 (0-5) | 0.008 |  |
| Tmax>8s, mL | 54 (22-103) | 67 (34-103) | 0.242 |  |
| Tmax>4s, mL | 171 (115-257) | 180 (129-255) | 0.969 |  |
| CBF<20%, mL | 0 (0-0) | 0 (0-7) | 0.028 |  |
| CBF<34%, mL | 5 (0-30) | 14 (0-30) | 0.028 |  |
| CBF<38%, mL | 8 (0-34) | 20 (5-43) | 0.014 |  |
| CBV<34%, mL | 4 (0-33) | 9 (0-36) | 0.170 |  |
| CBV<38%, mL | 5 (0-37) | 11 (0-39) | 0.137 |  |

The time from symptom onset to arterial puncture was comparable between the two groups. Outcome characteristics, however, revealed a higher mortality rate in the ICAD group compared to the non-ICAD group. This may partly be attributed to the higher proportion of patients with basilar artery occlusions in the ICAD group (Table 3).

**Table. 3.** Outcome characteristics.

**Outcome characteristics Table. 3**
|  | ICAD | Non-ICAD | P-Value |
| --- | --- | --- | --- |
|  | N=107 | N=119 |  |
| Hemorrhage | 17 (16) | 21 (19) | 0.576 |
| Symptomatic hemorrhage | 4 (4) | 8 (7) | 0.193 |
| mRS: 0-2 | 77 (72) | 89 (80) | 0.195 |
| Mortality | 16 (15) | 2 (2) | **<0.001** |

The 90-day mRS distribution stratified by the presence or absence of ICAD is shown in (Figure 1). Among ICAD patients, 72% achieved an mRS of 0–2, while 15% had an mRS of 6. In contrast, 80% of non-ICAD patients achieved an mRS of 0–2, and only 2% had an mRS of 6. Overall, patients without ICAD tended to have better outcomes, with a higher proportion achieving lower mRS scores (indicating less disability, though not significantly) and fewer deaths compared to ICAD patients.

**Figure:1.**
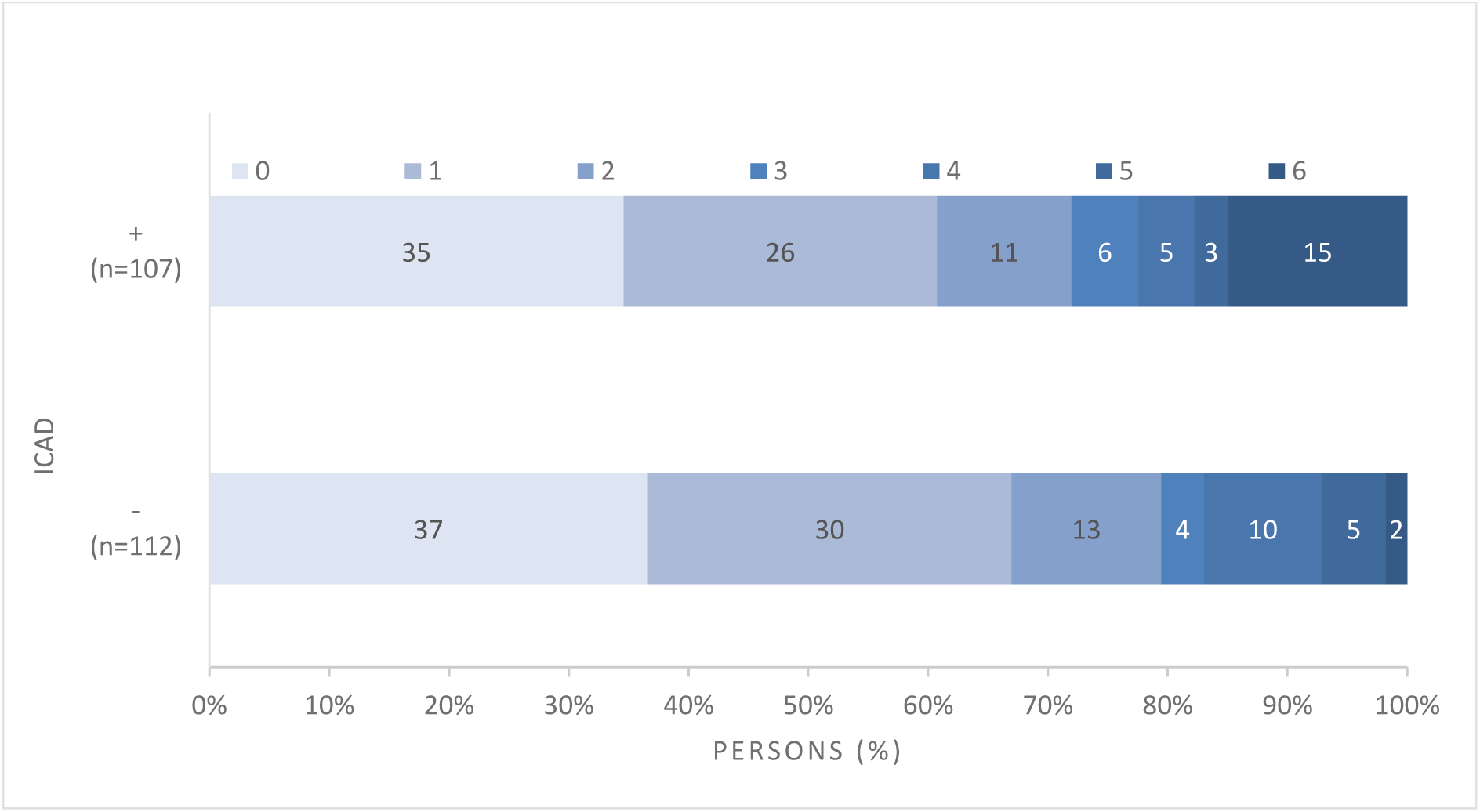
90-day mRS distribution stratified ICAD vs Non ICAD.

ROC curve analysis was performed for HIR to discriminate between ICAD and non-ICAD patients. HIR demonstrated modest diagnostic performance, with an area under the curve (AUC) of 0.62 (95% CI: 0.55–0.70; *p*=0.001). The optimal HIR cutoff was identified as 0.45, yielding a sensitivity of 0.66 and a specificity of 0.57. The test’s positive predictive value (PPV) was 0.60, and its negative predictive value (NPV) was 0.64, suggesting limited usefulness in distinguishing between ICAD and non-ICAD patients (Figure 2).

**Figure 2:**
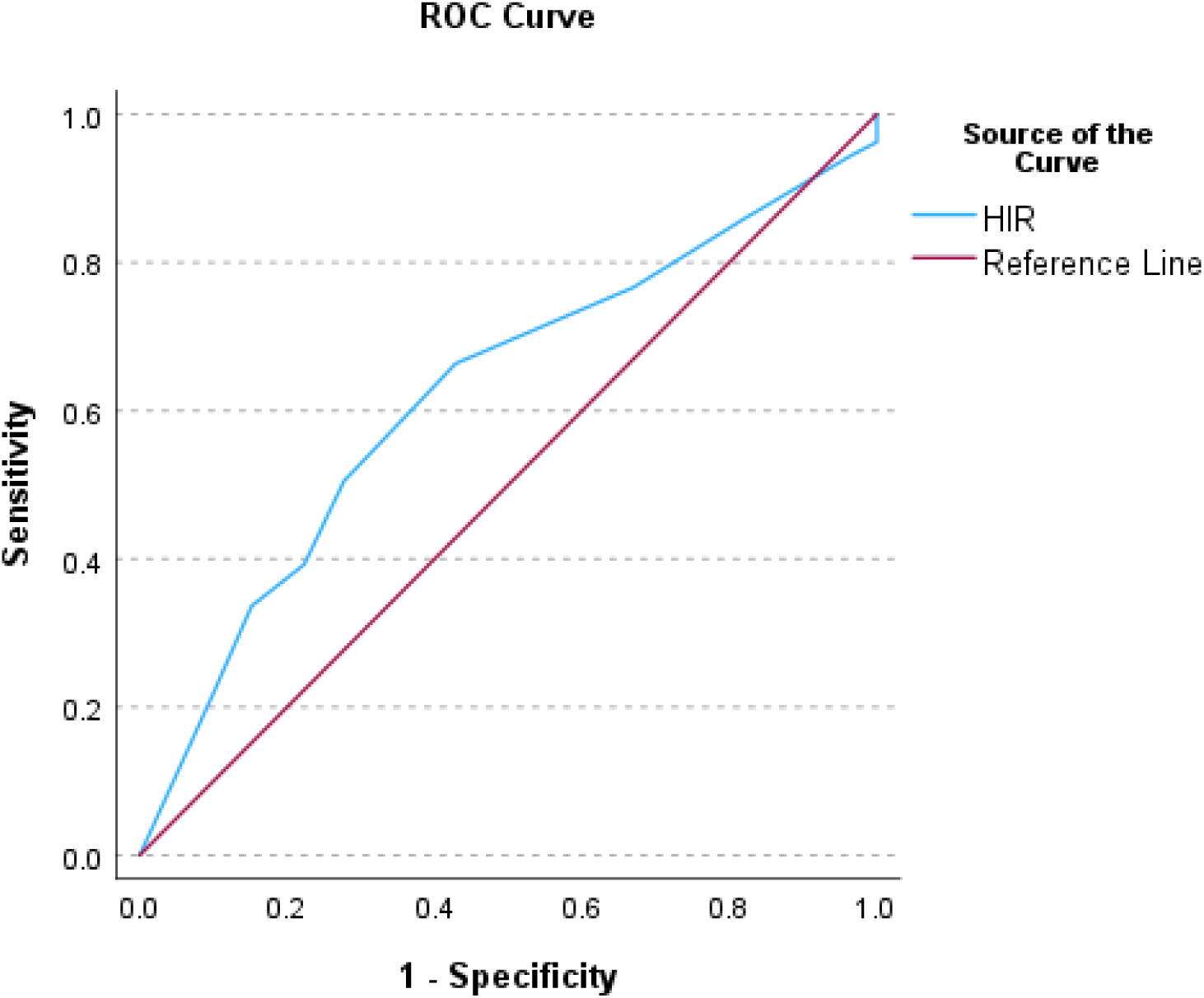
ROC curve analysis to discriminate ICAD using HIR.

For the CBV index, the ROC curve yielded an AUC of 0.57 (95% CI: 0.49–0.65; *p*=0.074). The optimal cutoff value was identified as 0.95, with a sensitivity of 0.20, specificity of 0.95, PPV of 0.78, and NPV of 0.55. This indicates limited sensitivity but high specificity for discriminating between ICAD and non-ICAD patients (Figure 3).

**Figure 3:**
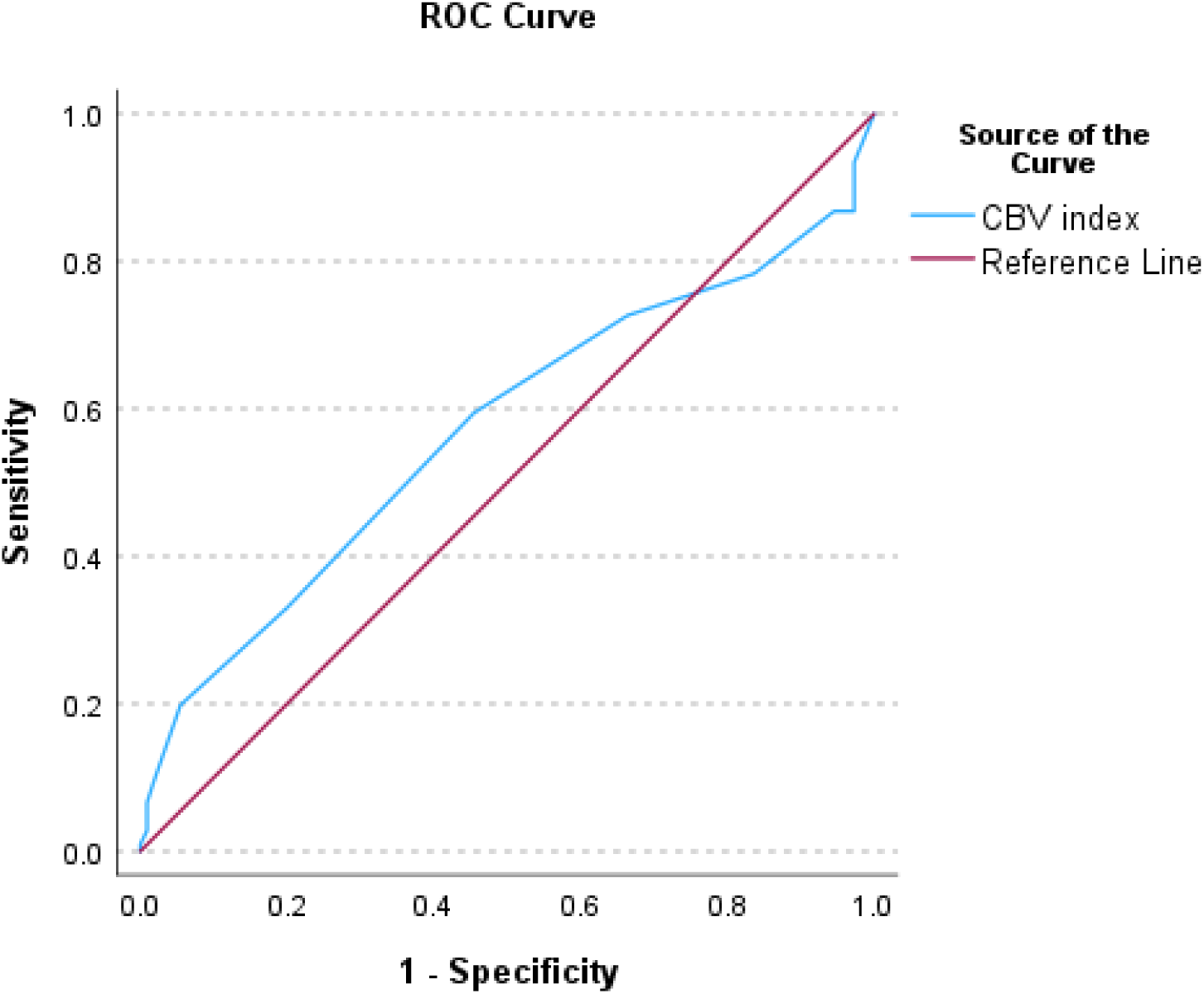
ROC curve analysis to discriminate ICAD using CBV index.

In multivariable analysis, the presence of ICAD, basilar occlusion, and larger core volumes were independently associated with increased mortality.

## Discussion

Our study demonstrates significant differences in risk factors, imaging profiles, clinical features, and outcomes between the ICAD and non-ICAD groups, particularly in the context of endovascular treatment (EVT) following interhospital transfer.

Patients with ICAD were older and exhibited a higher prevalence of vascular risk factors, including hypertension, diabetes mellitus, and smoking, which are well-documented contributors to ICAD in the literature. These findings align with previous studies, such as those by Al Kasab et al. ^(11)^, which identified similar comorbidities among ICAD patients. Importantly, ICAD patients in our cohort also had a higher incidence of prior stroke, emphasizing the recurrent nature of stroke in this population.

Our study showed that ICAD patients exhibited smaller core lesion volumes, higher mismatch ratios, and elevated HIR compared to non-ICAD patients. These findings suggest that patients with ICAD may have more preserved collateral circulation, which has been shown to correlate with better outcomes in acute ischemic stroke. Previous research by Imaoka et al. ^(12)^ supports the role of perfusion imaging markers, such as HIR, in distinguishing between ICAD and non-ICAD cases. Conversely, Wouters et al. ^(13)^ reported that fast infarct growth in non-ICAD LVO patients during transfer was associated with larger core volumes and poorer collateral circulation, highlighting the importance of early intervention.

The principal pathophysiology of ICAD is related to the buildup of cholesterol deposition within the intracranial arteries, leading to progressive intimal thickening and the formation of atheromatous plaques ^(6)(7)^. These plaques can result in stroke through mechanisms such as in situ thrombosis, artery-to-artery embolism, hypoperfusion, branch occlusion (lacune-like), or a combination of these processes. Many such lesions remain asymptomatic and are often detected incidentally during imaging for unrelated causes. ^(8)^

Our data indicate that ICAD patients had worse functional outcomes, as evidenced by higher modified Rankin Scale (mRS) scores at 90 days. While 72% of ICAD patients achieved an mRS of 0–2, the overall distribution of mRS scores suggested that non-ICAD patients had a better prognosis. This finding aligns with prior studies demonstrating that ICAD patients are less likely to achieve favorable functional recovery following EVT ^(13)^. The higher mortality observed in the ICAD group in our study was likely related to the greater number of patients with basilar artery occlusion. The higher mortality rate may also reflect the more complex nature of ICAD-related occlusions, which often involve multiple vessels and exhibit greater resistance to reperfusion therapies. Multiple studies have shown that patients with basilar occlusion tend to have worse prognoses. ^(14)^

The use of HIR and cerebral blood volume (CBV) index in our study provided valuable insights into the hemodynamic differences between ICAD and non-ICAD patients. However, while the HIR demonstrated modest diagnostic performance in discriminating between the two groups, its limited specificity and sensitivity suggest that it should be used in conjunction with other imaging parameters. This finding is consistent with earlier studies emphasizing the importance of a multimodal imaging approach for evaluating ICAD patients. ^(15)^

The limitations of our study include the retrospective design, which may introduce selection bias, as patients with more severe strokes were likely to undergo EVT. Additionally, the sample size was relatively small, and the study was conducted at a single comprehensive stroke center, which may limit the generalizability of the findings to other populations.

In conclusion, this study highlights the unique clinical and imaging characteristics of ICAD patients undergoing EVT. Despite more favorable imaging profiles, ICAD patients are at higher risk for mortality and worse functional outcomes, partly due to the increased prevalence of basilar occlusions. Future studies should focus on optimizing EVT strategies and exploring adjunctive therapies to improve outcomes in this challenging population.

## Data Availability

All data that support the findings of this article are available within the manuscript and its cited articles. Further inquiries can be directed to the corresponding author

## Acknowledgments

We sincerely thank all coauthors for their valuable time, effort, and contributions to this work.

## Sources of funding

No funding was received toward this work.

## Disclosures

None.

---

**Supplementary Table 1.**

|  | ICAD | Non-ICAD | p-value | Difference between medians (95% CI) |
| --- | --- | --- | --- | --- |
|  | N=107 | N=112 |  |  |
| Weight, kg | 76 (64-86) | 76.5 (67.8-86.1) | 0.603 |  |
| Temperature, °C | 36.7 (36.6-36.9) | 36.7 (36.6-36.9) | 0.814 |  |
| Hgb, g/L | 135 (118-152) | 135 (118-148) | 0.821 |  |
| PLT, 10<sup>9</sup>/L | 242 (193-315) | 242 (201-298) | 0.740 |  |
| HCT | 0.423 (0.370-0.456) | 0.417 (0.368-0.451) | 0.455 |  |
| PTT | 33.5 (31.4-36.5) | 34.9 (31.5-39.8) | 0.050 | -1.6 (-3.2-0) |
| INR | 1.1 (1.0-1.2) | 1.15 (1.1-1.3) | 0.001 | -0.1 (-0.1-0) |
| HbA1c | 7.1 (5.8-8.8) | 5.7 (5.1-7.0) | <0.001 | 1.0 (0.5-1.5) |
| Creatinine | 74 (61-90) | 75 (63-91) | 0.729 |  |

## Notes

### Competing Interest Statement

The authors have declared no competing interest.

